# Immune and metabolic beneficial effects of Beta 1,3-1,6 glucans produced by two novel strains of Aureobasidium pullulans in healthy middle-aged Japanese men: An exploratory study

**DOI:** 10.1101/2021.08.05.21261640

**Authors:** Nobunao Ikewaki, Tohru Sonoda, Gene Kurosawa, Masaru Iwasaki, Vidyasagar Devaprasad Dedeepiya, Rajappa Senthilkumar, Senthilkumar Preethy, Samuel JK Abraham

## Abstract

**Background:** Imbalances in glucose and lipid metabolism in the background of a declining immune system, along with aging, make one prone to glucolipotoxicity-related diseases such as hepatic steatosis and to high risk of infection-related mortality, as with COVID-19, warranting a safe prophylactic measure to help regulate both metabolism and the immune system. Based on the beneficial effects of the AFO-202 strain of black yeast *Aureobasidium pullulans*-produced beta 1,3-1,6 glucan in balancing of blood glucose and immune enhancement, and that of the N-163 strain of the same species in lipid metabolism and immune modulation, in this pilot study, we have evaluated their specific benefits in healthy human subjects.

**Methods:** Sixteen healthy Japanese male volunteers (aged 40 to 60 years) took part in this clinical trial. They were divided into four groups (n = 4 each): Group I consumed AFO-202 beta glucan (2 sachets of 1 g each per day), IA for 35 days and IB for 21 days; Group II consumed a combination of AFO-202 beta glucan (2 sachets of 1 g each) and N-163 beta glucan (1 sachet of 15 g gel each per day), IIA for 35 days and IIB for 21 days. Investigations for immune stimulation, anti-glycaemic, and anti-cholesterolemia biomarkers were undertaken in all four groups.

**Results:** In terms of metabolic control of glucose, the decrease in HbA1C and glycated albumin (GA) was significantly better in Group I compared with the other groups. Immune enhancement in terms of a significant increase of eosinophils and monocytes and marginal decrease in D-dimer levels, decrease in neutrophil-to-lymphocyte ratio (NLR), with an increase in the lymphocyte-to-CRP ratio (LCR) and leukocyte-to-CRP ratio (LeCR) was observed in Group I. Regulation of lipids by decrease in total and LDL cholesterol was better in Group II, and immunomodulation of coagulation-associated and anti-inflammatory markers by a decrease of CD11b, serum ferritin, galectin-3, fibrinogen was profound in Group II.

**Conclusion:** *A. pullulans,* a polythermotolerant black yeast - produced AFO-202 beta glucan has balanced blood glucose with marginal immune enhancement in healthy individuals, which when combined with N-163 beta glucan, balanced the lipid profile and immunomodulation. This outcome warrants larger clinical trials to understand the mechanisms and explore the potentials of these safe food supplements in prevention and prophylaxis of diseases due to dysregulated glucose and lipid metabolism, such as fatty liver disease, and infections such as COVID-19 in which a balanced immune activation and immunomodulation are of utmost importance, besides their administration as an adjunct to existing therapeutic approaches of both communicable and non-communicable diseases.

## Introduction

Metabolism imbalance is a gradually occurring condition leading to diabetes, heart disease, stroke, etc., and the risk varies between populations based on their genetic predisposition, diet, lifestyle, and environmental influences [1]. By the time an individual is diagnosed with any lifestyle illness requiring medication, further prevention and deceleration of the pathogenesis is an uphill task. To address this, exercise, dietary modifications such as intake of foods with low glycaemic index or fats, and medications are advised which are temporary and not definitive solutions [1]. The disease onset starts with elevated glucose levels leading to glucotoxicity in predisposed individuals and in high-fat diet populations; lipotoxicity occurs, and they both synergistically derail the metabolic homeostasis skewed towards dysregulation, paving the way for gradual onset of a nidus for diseases in many organs [2]. The liver, being the metabolic centre of the body, often becomes the key target [2], and coronary artery and cerebral systemic coagulopathies [1] may also occur. Against this background, the immune system is not only affected by the process of aging but also prematurely negatively implicated by glucotoxicity and lipotoxicity, which leads to a loss of balance between the generation of reactive oxygen species (ROS) and the ability of the endogenous antioxidant system (AOS), in turn causing aging and various age-related chronic pathologies such as inflammation, neurodegenerative diseases, atherosclerosis, and vascular complications of diabetes mellitus [3]. Additionally, the immune reserves may be depleted in handling, and the circulating high levels of advanced glycation end products (AGEs) and lipids affects the functional capability of the immune cells, leading to high risk of disease severity [3] when COVID-19-like infections occur [4]. The fibrosis that ensues after a chronic inflammation-metabolic-immune dysregulation can lead to pulmonary or liver fibrosis, such as non-alcoholic steatohepatitis (NASH) [2], which could eventually culminate in carcinogenesis [5]. Glucotoxicity and lipotoxicity also cause gut dysbiosis [6], which is now increasingly considered the key factor influencing progression of infections, inflammations, and fibrosis, creating a viscous cycle.

Against the given background, as a remedy, what we require ideally should:

I. **Regarding glucotoxicity and lipotoxicity:**

1. ***At an early stage or before onset of disease:***
• Balance the blood glucose levels, especially the post-prandial spike
• Balance the blood cholesterol level without side effects
2. ***Post-disease onset stage:*** During and after onset of the glucotoxicity and/or lipotoxicity
• Control glucose and cholesterol with no interaction with other drugs prescribed
• Balance the blood cholesterol level without side effects; control LDL and VLDL without affecting HDL
• Be able to control inflammation and the accumulation of free fatty acids (FFA)
3. ***After progression of disease with chronic sequalae stage*:** Post-onset of pathogenesis causing organ and/or systemic inflammation
• Control organ inflammatory reaction to avoid fibrosis
• Balance micro-inflammation of the gut
**II. Regarding systemic wellness and immune balance:**

1. All through the above stages:
• Support the immune system, especially during aging, by enhancing it to prevent illnesses from disease-affected weakness
• Promote immune modulation to avoid hyper-activation and cytokine storm
• Balance immune enhancement and modulation to avoid pre-disposing factors to carcinogenesis.
• Reverse gut dysbiosis

Although a single such prophylactic measure or component is almost impossible, we selected two products of strains from the black yeast *A. pullulans* which have a track record of safety [7–9].

### AFO-202 benefits

The AFO-202 strain-produced beta glucan has been shown to normalize Hba1c and fasting, post-prandial blood glucose levels in patients with type II diabetes [7]. It has been shown to decrease elevated LDL and VLDL cholesterol and triglycerides in clinical studies of metabolic syndrome [8]. Enhancement of immune cells such as natural killer (NK) cells and macrophages, apart from suppression of pro-inflammatory cytokines such as IL-1beta, IL-2, IL-6, IL-12 (p70+40), interferon-gamma (IFN-gamma), tumour necrosis factor-alpha (TNF-alpha), or soluble Fas ligand (sFasL) while enhancing beneficial cytokines such as interleukin-8 (IL-8) or soluble Fas (sFas) and antibodies has been reported [9]. Apart from these beneficial immune and metabolic modulations, a decrease in the neutrophil-to-lymphocyte ratio (NLR) and increase in lymphocyte-to-C-reactive protein (CRP) ratio (LCR) and leukocyte-to-CRP ratio (LeCR) are particularly significant in COVID-19 [10], as the dysregulation of these parameters has been correlated with progression of the disease and higher odds of mortality [11]. The potential of the AFO-202 beta glucan as an immune adjuvant in the prophylaxis of COVID-19, along with beneficial anti-coagulopathy benefits, has been described [12–15].

### N-163 benefits

While AFO-202 is relevant to both metabolic and immune regulation, the anti-inflammatory, anti-fibrotic potential of N-163 has been reported with significance in a NASH animal model [16], along with a decrease in inflammation-associated lipid parameters such as non-esterified free fatty acids (NEFAs) [17]. Thus, N-163 is more relevant in the stages of progressed disease status.

Before addressing specific disease targets, we sought to study the effects of AFO-202 and N-163-produced beta glucans in the middle-aged, healthy subjects, as they have been the most vulnerable population for metabolic diseases [18] and severe COVID-19.

## Methods

The study was conducted in compliance with the ethical principles based on the Declaration of Helsinki and the Ethical Guidelines for Medical Research Involving Human Subjects (notified by the Ministry of Education, Culture, Sports, Science and Technology and the Ministry of Health, Labour and Welfare, Japan). The study protocol was approved by the institutional review board (IRB) of Chiyoda Paramedical Care Clinic, Tokyo, Japan (study protocol number GNC20C1), and registered with the University Hospital Medical Information Network-Clinical Trial Registry (UMIN-CTR) of Japan [19]. The study was conducted at the Chiyoda Paramedical Care Clinic, Tokyo, Japan.

### Study Subjects

The study was designed as an exploratory study in healthy Japanese male volunteers aged 40 to 60 years with four intervention conditions: two test food groups and two durations of intake in each test food group. As the minimum number of participants required for statistical comparisons within and between intervention conditions is four per intervention condition, a total of 16 target study participants was determined.

The person in charge of the allocation, as specified in the study protocol, allocated the study subjects to the four groups as evenly as possible, giving first priority to pre-test BMI, second priority to weight, and third priority to height.

Subjects who met the selection criteria in Table 1 and did not fall under any of the exclusion criteria were eligible for the study.

**Table 1.**
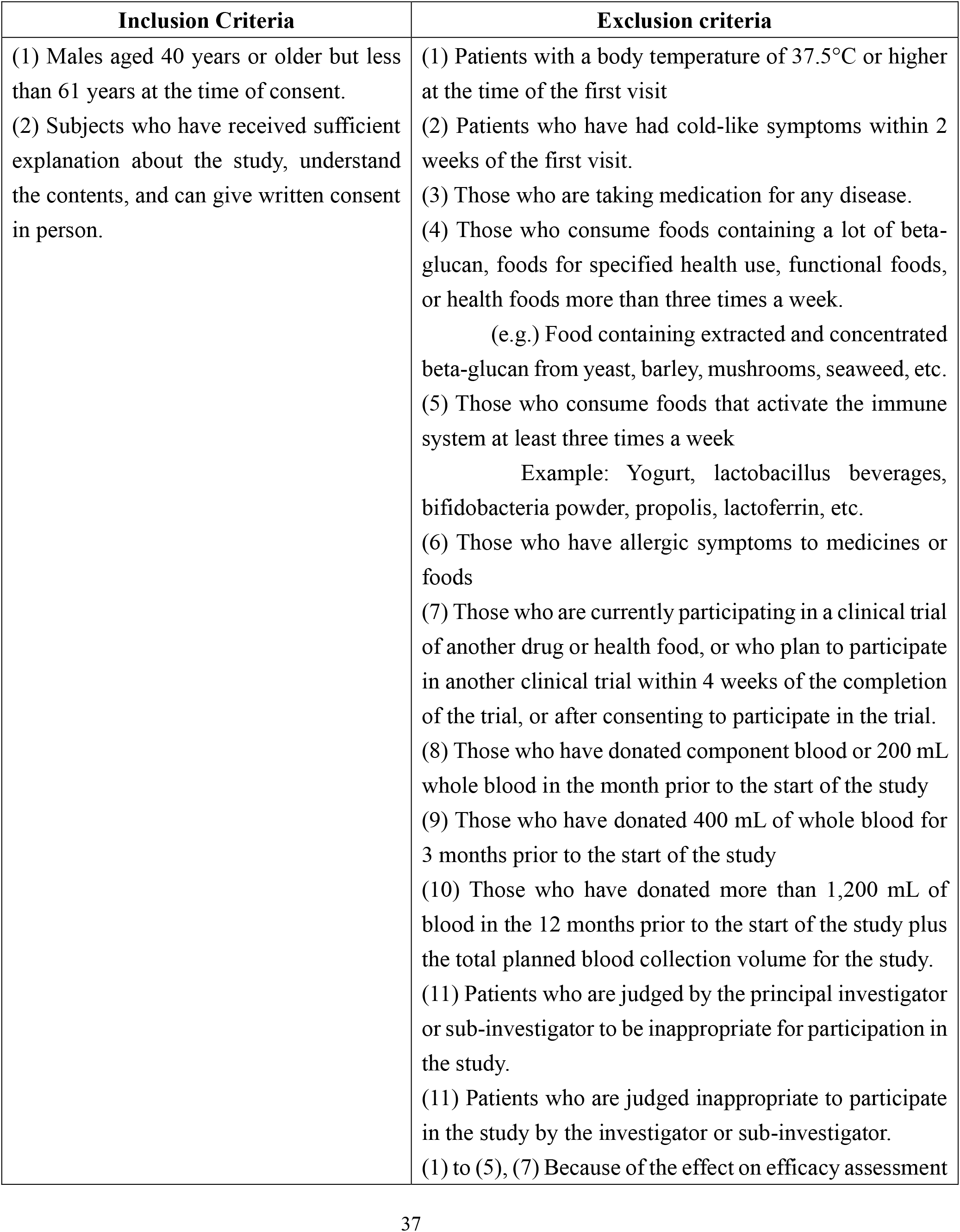
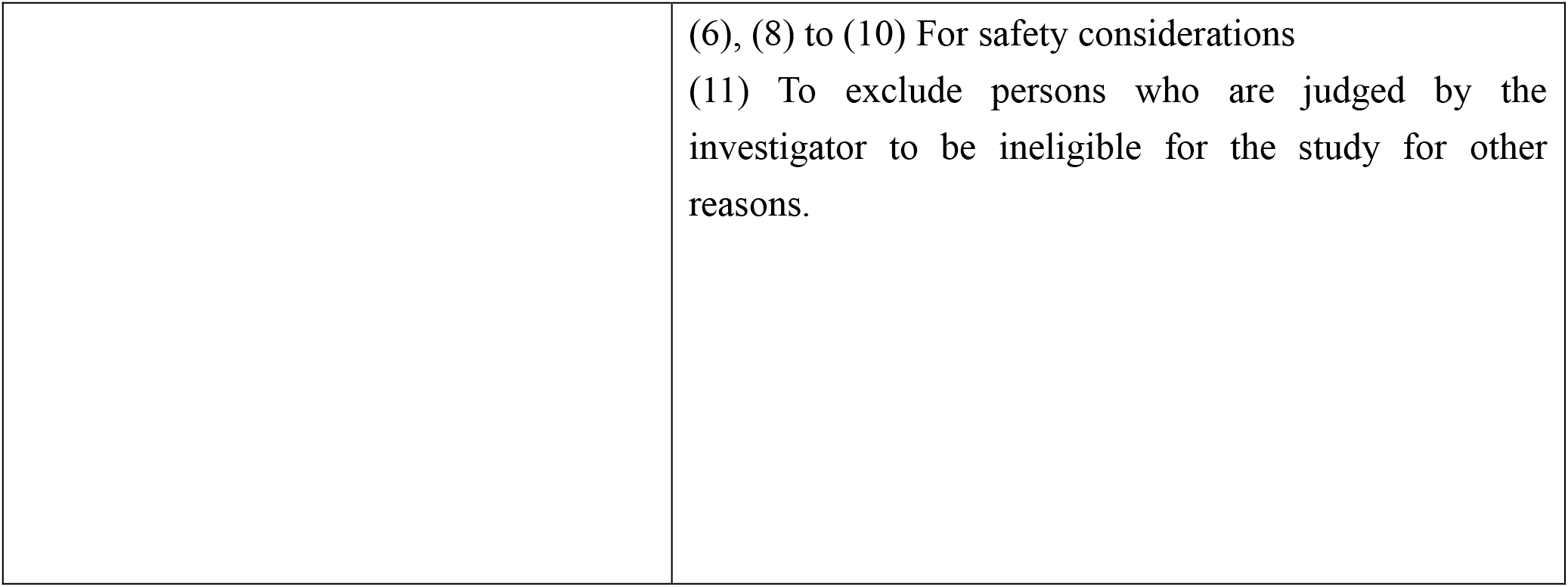
: **Inclusion and exclusion criteria**

### Intervention

The duration of the study food intake and the schedule of visits for each group are shown in Table 2.

**Table 2.**
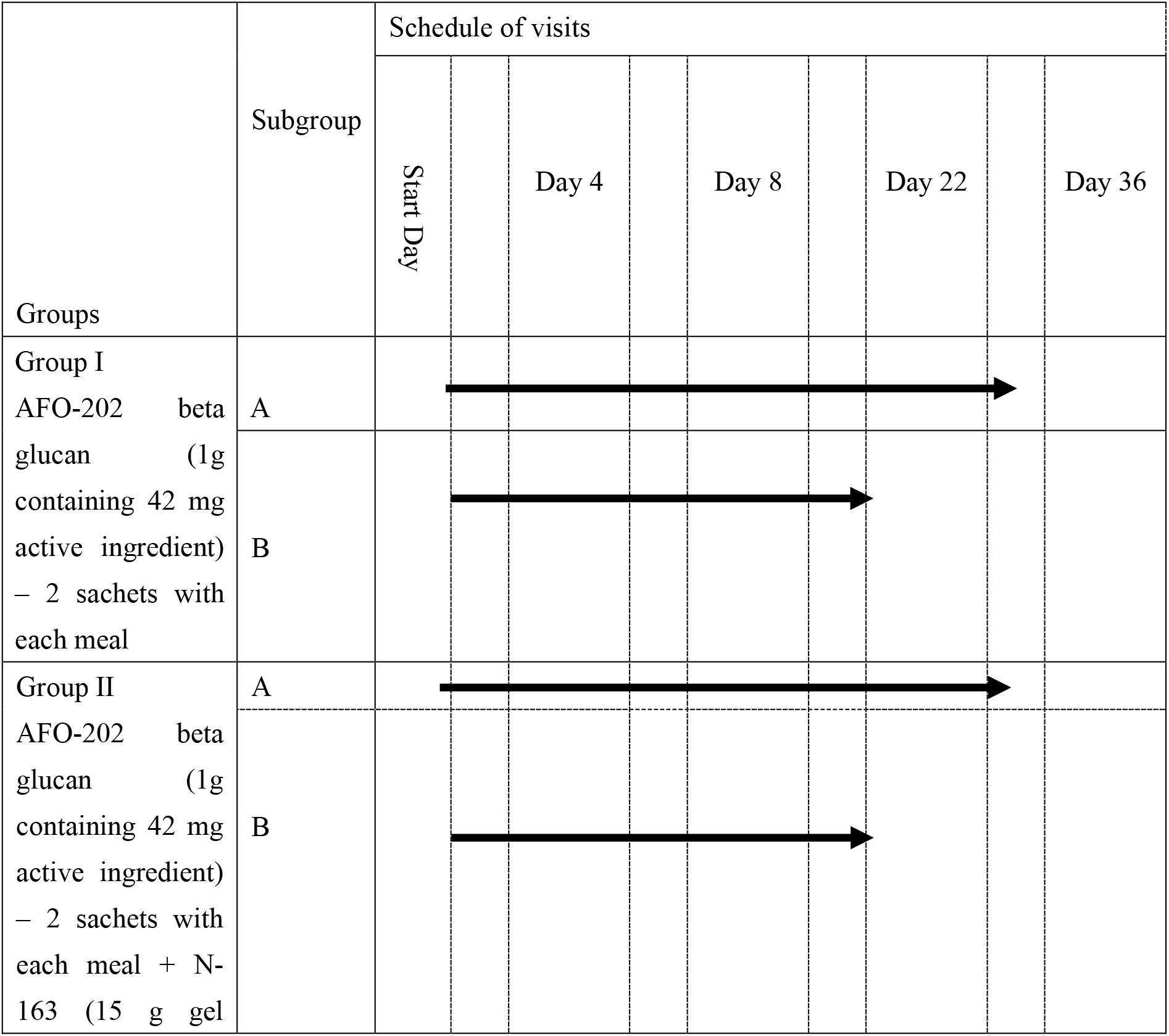
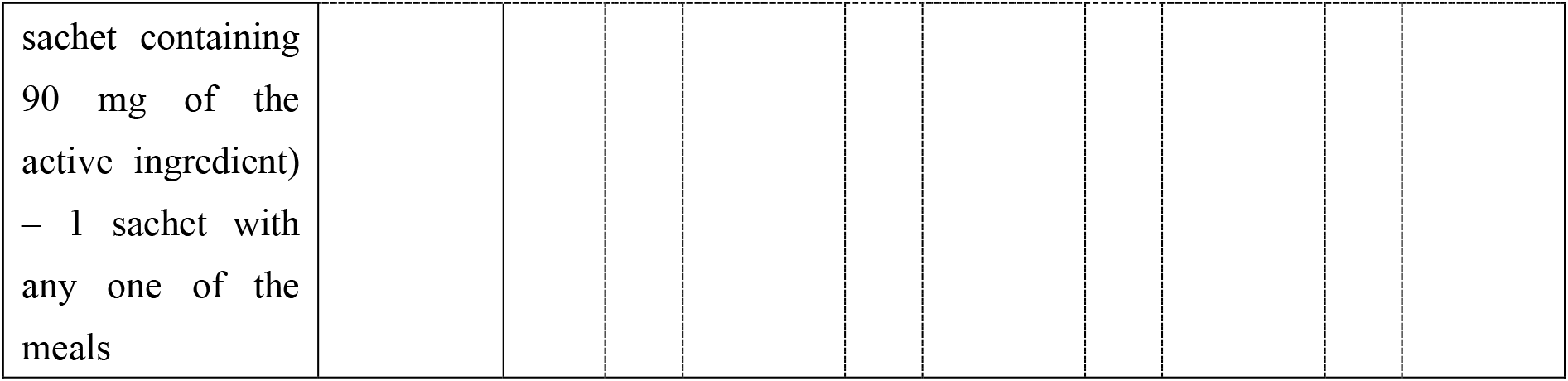
: **Interventions and Study groups**

## Evaluations

The following tests were carried out after written consent was obtained from the study subjects.

### Pre-test and before intake

Blood sampling volume: 34 mL

Background survey: gender, date of birth, age, smoking habits, drinking habits, eating habits, current medical history, medication, treatment, previous history, allergies (to drugs and food), regular use of food for specified health purposes, functional foods, health foods, intake of foods rich in β-glucan, intake of immunity-boosting foods, and blood donation (within 1 year).

At *pre-test and before intake,* Day 4 of intake, Day 8 of intake, Day 22 of intake, Day 36 of intake, and Day 15 of post-observation

➢ Medical history and physical measurements: medical history, height, weight, BMI, temperature
➢ Physiological examination: systolic blood pressure, diastolic blood pressure, pulse rate
➢ Haematology 1: White blood cell (WBC) count, red blood cell (RBC) count, haemoglobin (Hb), haematocrit (Ht), MCV, MCH, MCHC, platelet (PLT) count
➢ Haematology 2: Basophil, Eosinophil, Neutrophil, Lymphocyte, Monocyte counts
➢ Haematology 3: D-dimer, prothrombin time (PT), Ferritin, Fibrinogen
➢ Blood biochemistry: T-Cholesterol, LDL-Cholesterol, HDL-Cholesterol, triglycerides (TG), HbA1c (NGSP), glycated albumin (GA)
➢ Immunological test : CRP, blood IgG, blood IgM, blood IgA
➢ Cellular immunity test: CD11b in monocyte fraction
➢ IL-2, IL-6, IL-7, IL-8, IFN-γ, sFas ligand
➢ Galectin-3

### Daily diary

Daily diary: Participants kept a diary from the day of the start of test food consumption until the day before the 36th day of consumption and the 15th post-observation day.

The following items were recorded in the diary: intake of test foods, body temperature; intake of food for specified health uses, functional foods, and health foods; intake of restricted foods; subjective symptoms; visits to medical institutions; treatment; and use of medicines.

### Examples of restricted foods

The following are examples of restricted foods.

Supplements rich in beta-glucan: supplements containing beta-glucan extracted and concentrated from yeast, barley, mushrooms, and seaweed.

Foods claimed to stimulate the immune system: yoghurt, lactobacillus beverages, bifidobacteria powder, propolis, lactoferrin, etc.

### Primary endpoints

1. Immune activation effect
2. WBC, RBC, Hb, Ht, PLT, MCV, MCH, MCHC
3. Basophils, eosinophils, neutrophils, lymphocytes, monocyte counts
4. CRP, IgG in blood, IgM in blood, IgA in blood
5. IL-2, IL-6, IL-7, IL-8, IFN-γ, sFas ligand

### Secondary endpoints

1. Coagulopathy related markers

**i.** Ferritin, D-dimer, PT, Fib, CD11b in monocyte fraction, galectin-3
2. Blood glucose level
3. HbA1c, GA
4. Cholesterol level

**i.** TG, T-Cho, HDL-Cho, LDL-Cho

### Safety evaluation items

Incidence of adverse effects

## Data analysis

### Target population for analysis

After all data had been obtained, a decision was made on the handling of cases and the acceptance or rejection of data for all cases. Intention to treat (ITT) was defined as the group of all study subjects excluding those who did not consume the study food. The Full Analysis Set (FAS) was defined as the population from which the ITT excluded subjects who withdrew or dropped out of the study. The FAS excluding subjects who met the following exclusion criteria for PPS analysis was defined as the Per Protocol Set (PPS).

### Exclusion criteria for PPS analysis

All study subjects who consumed the test food were included in the analysis. However, if a study subject met any of the following exclusion criteria, the study investigator and sponsor discussed and decided how to handle the test results.

1) Less than 85% intake of the study food
2) Less than 85% of the diary entries made
3) Repeated failure to take the indicated precautions
4) Failure to comply with the fasting and smoking cessation rules, or any other significant behaviour that undermined the reliability of the test results
5) A breach of the exclusion criteria determined after completion of the test
6) Other obvious reasons for omission

### Data-handling criteria

For each test item in the relevant test, if the measured value could not be obtained because it was below the lower or above the upper limit of measurement, the lower or upper limit of measurement was substituted. Values exceeding the range of three times the standard deviation from the mean value for each test item were considered outliers and were not used in the analysis. Missing values were not complemented, but there were no missing values in this study.

### Statistical analysis

The statistical significance level was set at 5%, two-sided. SPSS26.0 (IBM Japan, Ltd.) and Microsoft Excel (Microsoft Corporation) were used as analysis software. An unpaired t-test, Fisher’s exact test (Bonferroni correction), Dunnett certification, and a correspondence t-test were performed.

## Results

Sixteen patients (ITT) who fulfilled all the selection criteria and none of the exclusion criteria were selected to start the study. Because one study subject (No. 4) with leukocyte abnormalities (suspected leukaemia) discontinued or dropped out of the study, 15 study subjects who participated throughout the entire study period were considered FAS. In addition, two study subjects (Nos. 11 and 16) were excluded as a result of deliberation at the case review meeting because they fell under “6) Other obvious reasons for omission” in the “*Exclusion criteria for PPS* analysis” section. The reason for the exclusion of study subject No. 11 was that there were more outliers (mean ± 3SD) for this subject than for other study subjects, and it was judged that the exclusion might affect the study results. Thus, subjects Nos. 11 and 16 were excluded from the analysis. After excluding these two subjects from the FAS, 13 subjects were included in the PPS. The CONSORT diagram of the trial is presented as figure 1. There were no significant differences between the test food groups (four groups) in terms of BMI, height, and weight, which were the test subject allocation factors.

**Figure 1:**
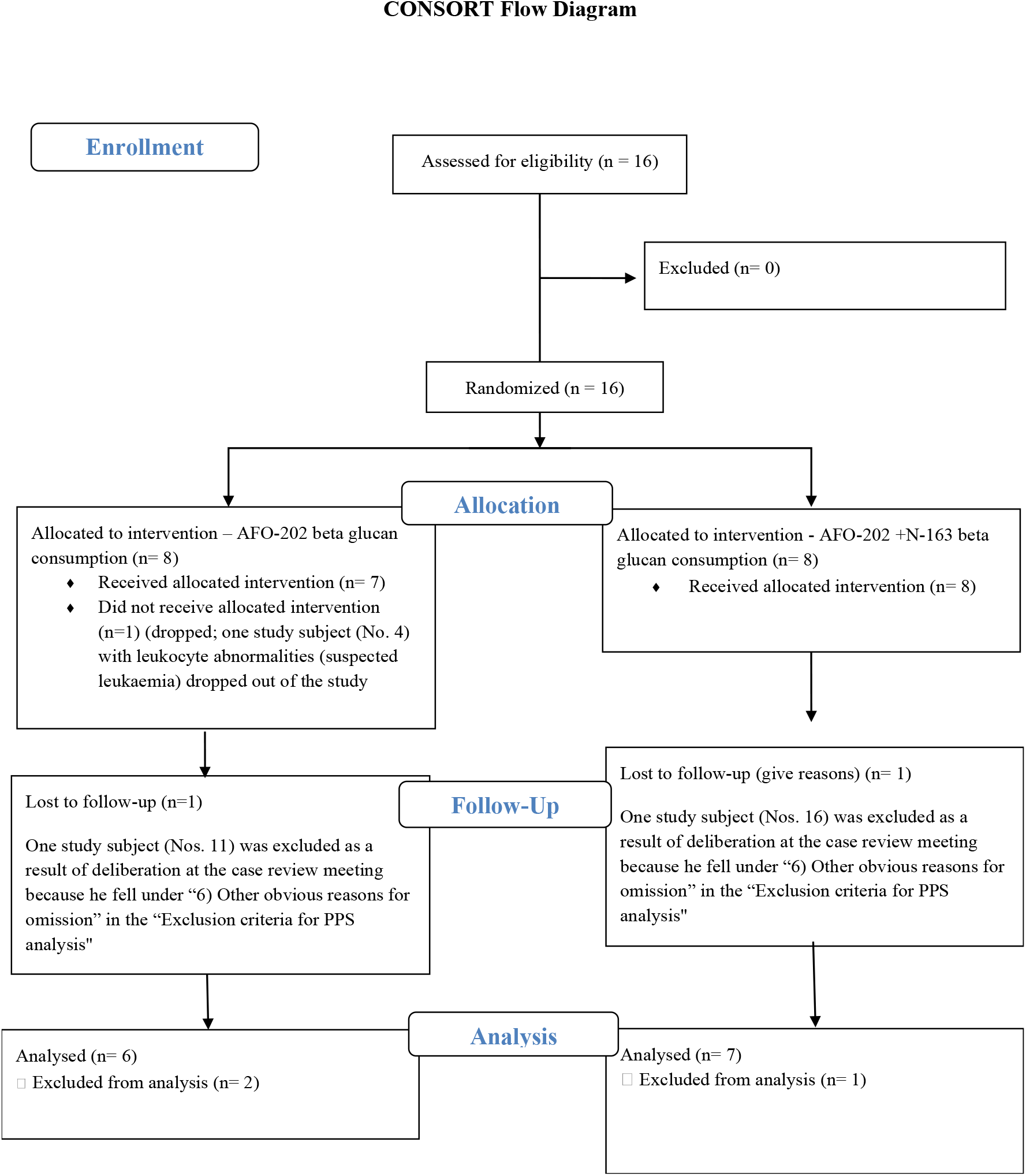
CONSORT Flow diagram of the trial

The intake rates (number of days and number of packets) for FAS and PPS were 100.00% for both test food groups (I and II).

Comparisons between the test food groups using the change from pre-consumption values showed statistically significant differences in the parameters outlined in Table 3.

I. **AFO-202 beta glucan:**

1. ***Glucose metabolism:***
*HbA1C:*
In Group I, the decrease was greater by -0.23 ± 0.06% after 35 days of intake compared with Group II (-0.08 ± 0.05%), which showed a statistically significant higher value (p < 0.05) (Figure 2A).
*Glycated albumin (GA):*
After 21 days of consumption, the GA decrease in Group I (-0.53 ± 0.15%) was statistically significantly higher than that of the Group II (-0.10 ± 0.18%) (p < 0.05), (Figure 2B).
2. ***Haematological indices of immune stimulation:***
*RBC*
After 21 days of consumption, the RBC was statistically significantly higher (p < 0.05) in Group I (4.0 ± 5.3 x 104/µL) compared with test Group II (-8.8 ±5.6 x 104/µL).
*Hb*
After 21 days of consumption, the value in Group I (0.13 ± 0.12 g/dL) (p < 0.01) was statistically significantly higher compared with that of Group II (-0.38 ± 0.15 g/dL).
*Haematocrit (Ht)*
After 21 days of intake, Group I (-0.03 ± 0.40%) showed statistically significant higher Ht values than did Group II (-1.50 ± 0.29%) (p < 0.01).
*Eosinophils*
A statistically significant difference was found between the test food groups in terms of the change from pre-treatment to post-treatment (p < 0.05). Eosinophil count (0.50 ± 0.54%) was higher in Group I compared with Group II (-0.36 ± 0.61%) (Figure 3A).
*Monocytes*
After 7 days of consumption, Group I (6.63 ± 0.51%) showed a statistically significantly higher monocyte value than did Group II (5.00 ± 0.82%) (p < 0.05). After 21 days of consumption, Group I (1.93 ± 0.47%) also showed a statistically significant increase compared with Group 2 (0.87 ± 0.21%) (p < 0.05) (Figure 3B).
*CRP*
At 21 days, the decrease in CRP was greater in Group I (level= 0.0517 mg/dl) compared with Group II (0.1329 mg/dl), which was statistically significant (p < 0.05) (Figure 3C).
*IL-7*
After 7 days of consumption, the IL-7 level was statistically significantly higher (p < 0.05) in Group I (4.33 ±0.87 pg/mL) compared with Group II (2.67 ±0.55 pg/mL).
*IL-8*
Group I’s IL-8 values (7.003 ±0.929 pg/mL) were statistically significantly higher than those of Group II (5.230 ±0.469 pg/mL) after 7 days of intake (p < 0.05).
*D-dimer*
After 35 days of intake, the D-dimer decrease in Group I (-0.30 ±0.10 µg/mL) was statistically significantly higher than that of the test food Group II (0.00 ±0.10 µg/mL) (p < 0.05) (Figure 3D).
*NLR, LCR, and LeCR*
The decrease in NLR was greater in Group I at day 21, but at day 35, the decrease was higher in Group II. In terms of LCR and LeCR, at day 35, the increase from baseline value was greater in Group I compared with Group II (Figure 4A-C). The results however were not statistically significant.
II. **N-163:**

1. **Regulation of lipid parameters:**
*Total cholesterol (T-Cho):*
After 21 days of intake, the T-Cho decrease in Group II (-12.8 ± 4.0 mg/dL) was statistically significantly higher than that of the test food Group I (9.0 ± 12.3 mg/dL) (p < 0.05) (Figure 5A).
*LDL cholesterol (LDL-Cho)*
There was a statistically significant decrease in LDL-Cho in Group II, at 124.0 ±25.3 mg/dL, after 21 days of consumption, compared with 134.0 ±25.2 mg/dL before consumption (p < 0.01) (Figure 5B).
2. **Immuno-modulation and anti-inflammatory effects:**
*IL-2*
The increase to 0.3743 ± 0.1165 pg/mL after 14 days of post-observation in Group II was statistically significant higher (p < 0.05) than the 0.1220 ± 0.0635 pg/mL value in Group 1.
*Blood IgA*
After 21 days of intake, Group II (340.3 ±64.9 mg/dL) had a statistically significantly higher blood IgA value than did Group I (175.0 ±9.5 mg/dL) (p < 0.01).
*MCHC*
After 7 days of consumption, the MCHC was statistically significantly higher (p < 0.05) in Group II (32.56 ± 0.55%) compared with Group I (31.85 ± 0.55%).
*Serum galectin, ferritin, and fibrinogen*
The decrease in serum fibrinogen, ferritin and galectin-3 was greater in Group II compared with Group I, but the difference was not significant (Figure 6A-C).
*CD11b*
An increase in CD11b in the monocyte fraction was observed in Group II after 21 days of ingestion compared with Group I but it was not statistically significant (Figure 6D).
*Other parameters:*
No statistically significant difference was observed in the other parameters.
*Safety endpoints (incidence of adverse reactions):*
No adverse reactions occurred in this study.

**Figure 2:**
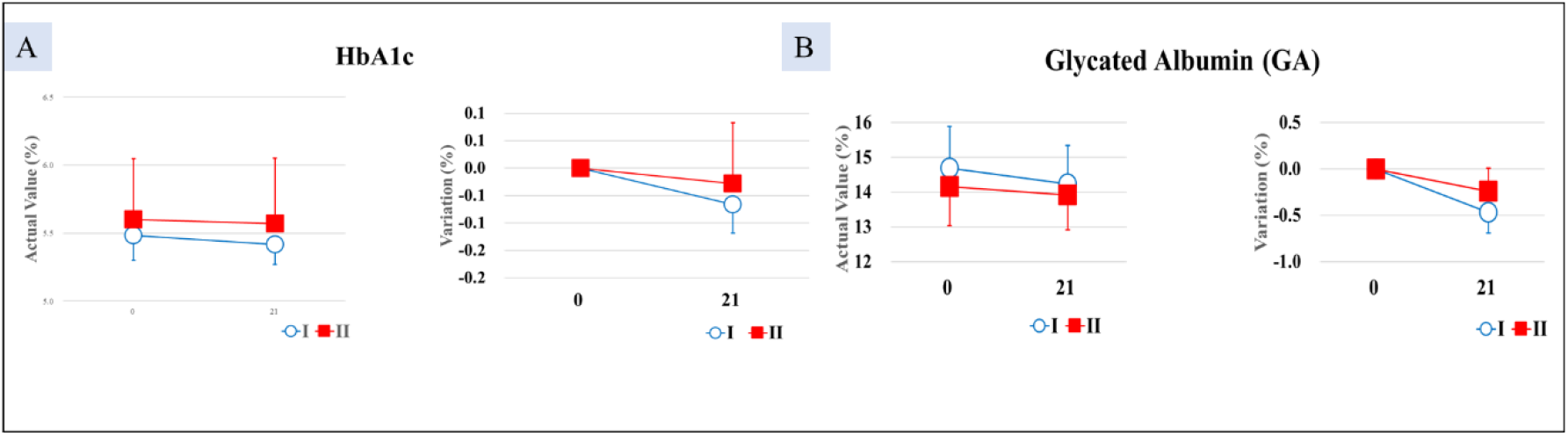
Decrease in A. HbA1c; B. Glycated albumin (GA); significantly greater in Group I (AFO-202 beta glucan) compared to Group II (AFO-202+N-163 beta glucan)

**Figure 3:**
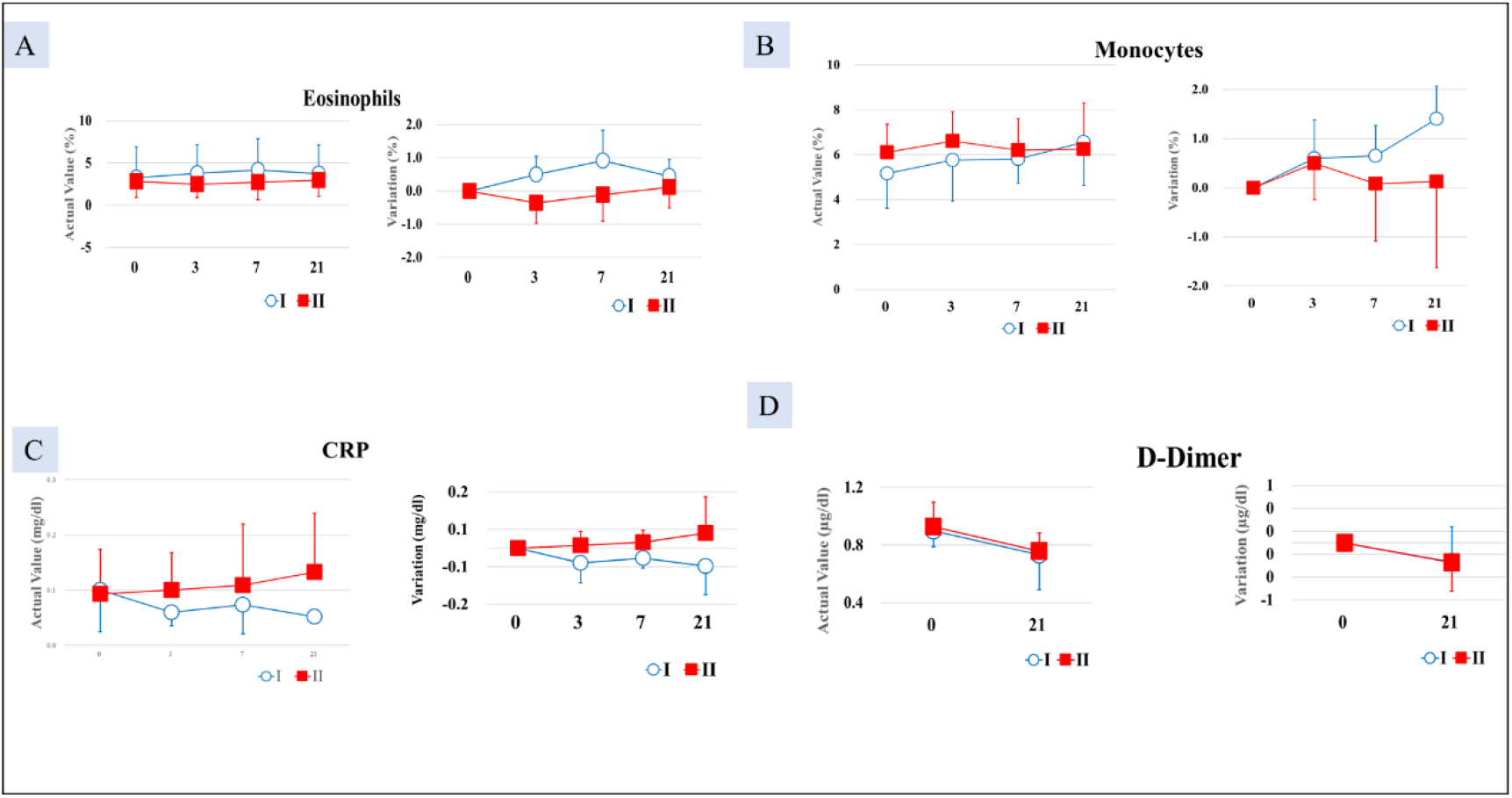
Increase in A. Eosinophil count; B. Monocytes count significantly greater in Group I (AFO-202 beta glucan) compared to Group II (AFO-202+N-163 beta glucan); decrease in C. CRP and D. D-Dimer, significantly greater in Group I (AFO-202 beta glucan) compared to Group II (AFO-202+N-163 beta glucan)

**Figure 4:**
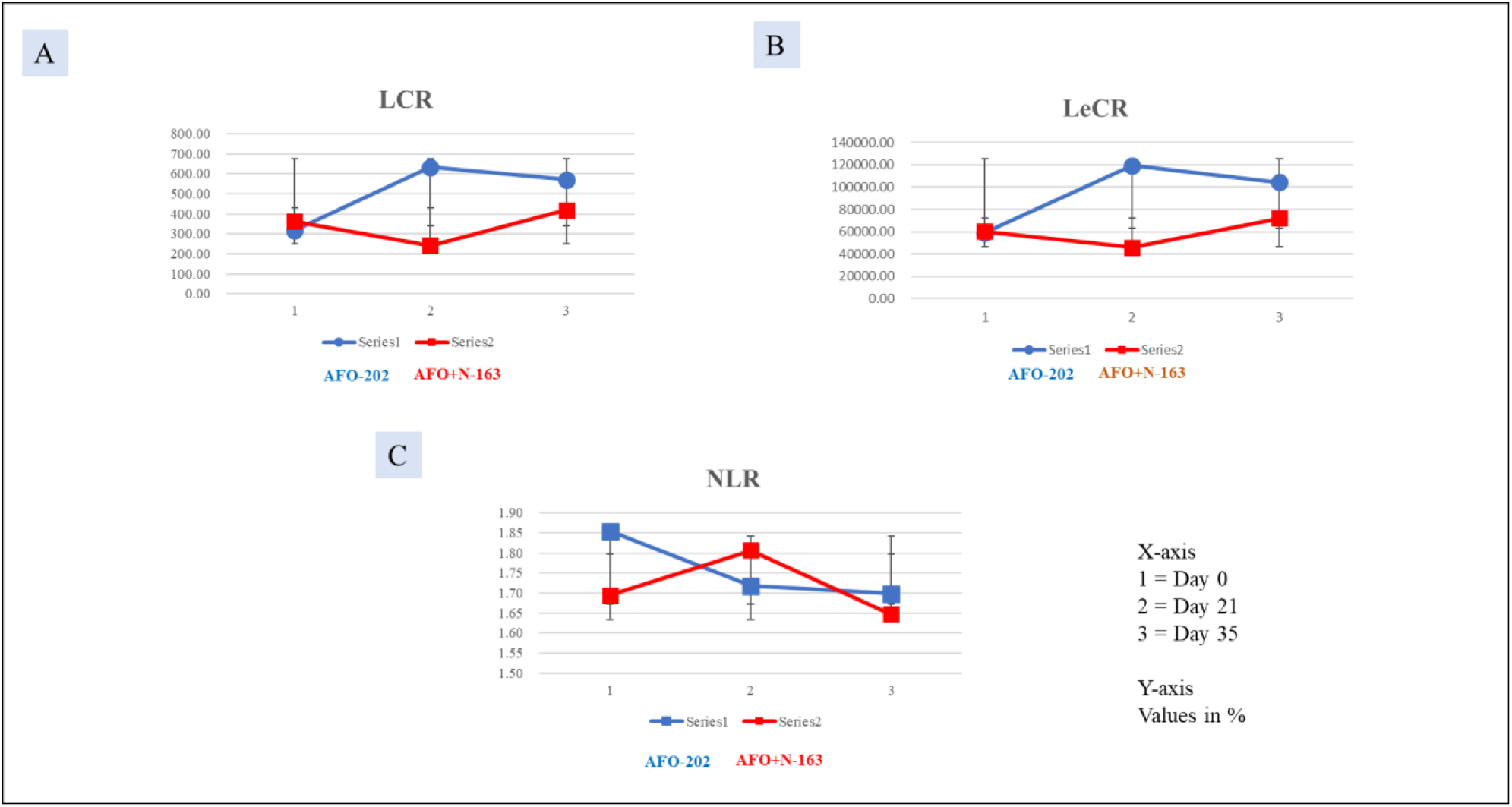
Increase in A.LCR; B. LeCR and decrease in C. NLR significantly greater in Group I (AFO-202 beta glucan) compared to Group II (AFO-202+N-163 beta glucan)

**Figure 5:**
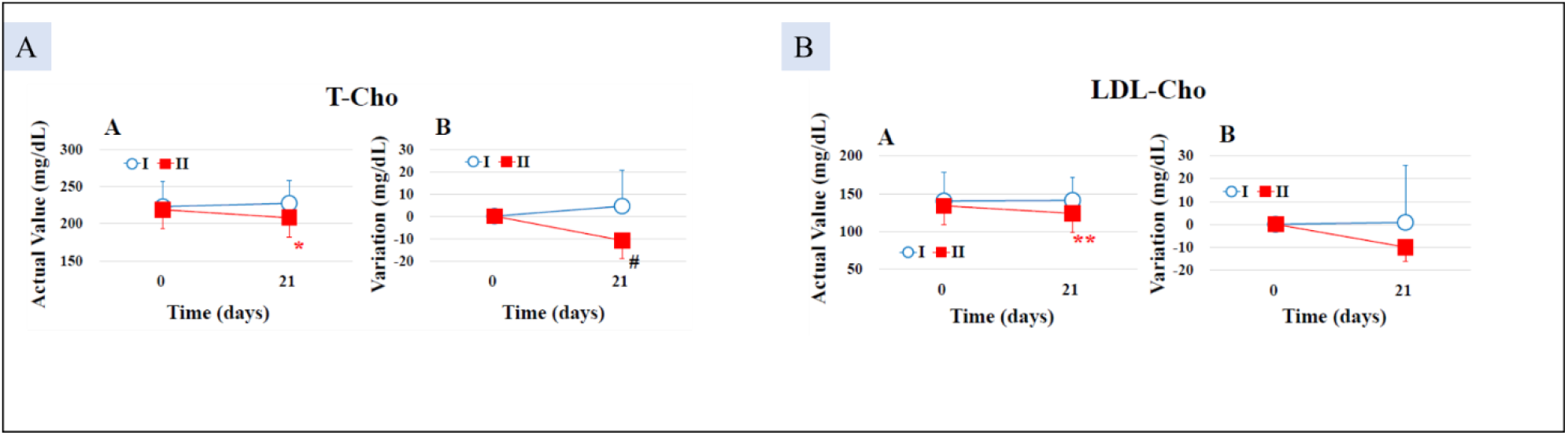
Decrease in A. total cholesterol (T-Cho) and; B. LDL-cholesterol significantly greater in Group II (AFO-202+N-163 beta glucan) compared to Group I (AFO-202 beta glucan)

**Figure 6:**
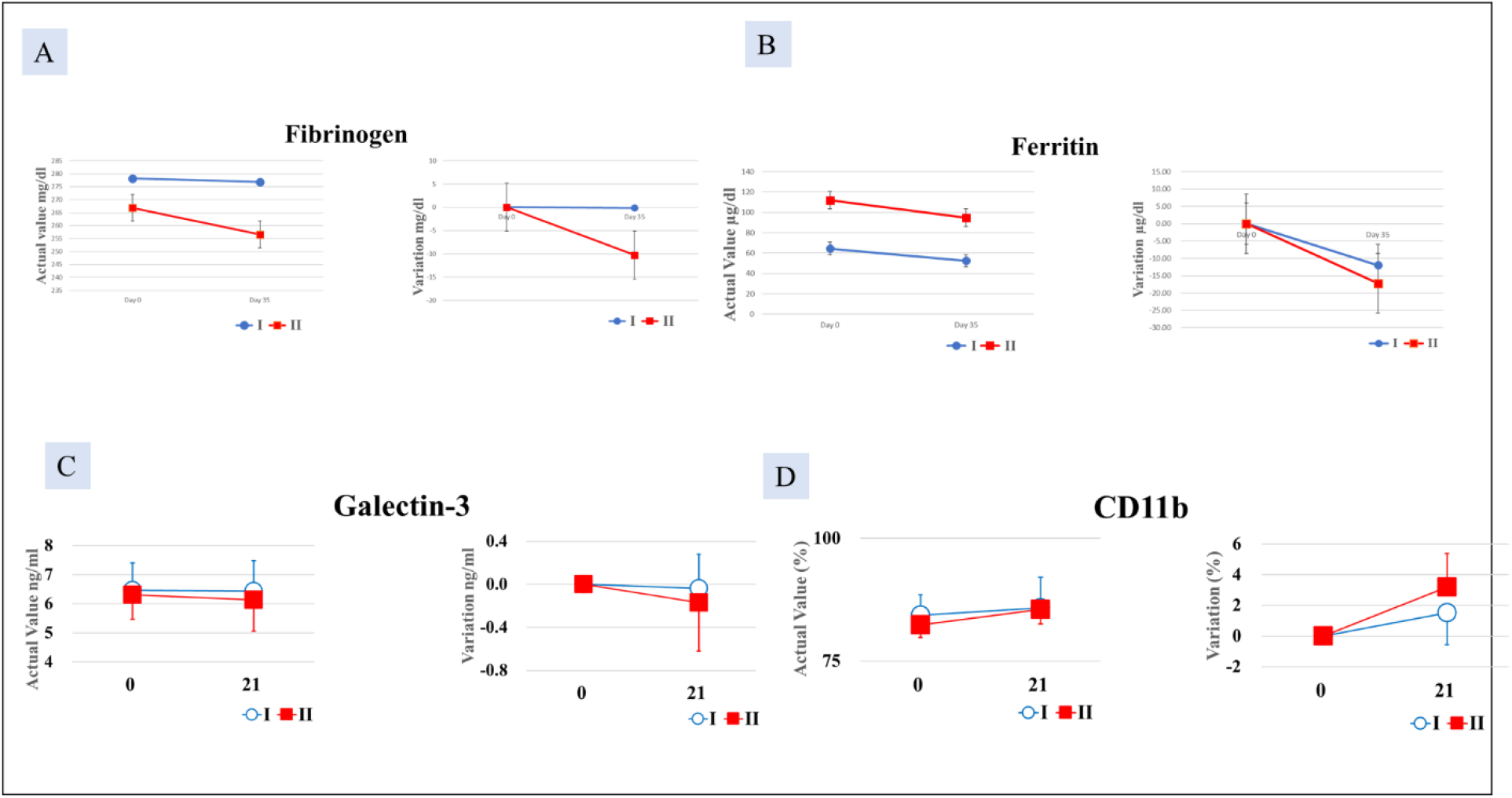
Decrease in A. Fibrinogen and; B. Ferritin C. Galectin-3 and D. increase in CD11b greater in Group II (AFO-202+N-163 beta glucan) compared to Group I (AFO-202 beta glucan)

**Table 3:** AFO-202 vs AFO-202+N-163 Beta glucans in healthy volunteers; results in a nutshell.

| Beneficial effect | Biomarker | Outcome | Effective compound | P Value significance |
| --- | --- | --- | --- | --- |
| **Immune enhancement** | Monocyte | **Increase** | AFO-202 | <0.05 |
|  | Eosinophils |  | AFO-202 | <0.05 |
|  | CRP |  | AFO-202 |  |
|  | IL-7 |  | AFO-202 | <0.05 |
|  | IL8 |  | AFO-202 | <0.05 |
| **Immunomodulation** | IgA | **Increase** | AFO-202 +N-163 | <0.01 |
|  | IL-2 |  | AFO-202 +N-163 | <0.05 |
|  | LCR |  | AFO-202 +N-163 |  |
|  | LeCR |  | AFO-202 +N-163 |  |
|  | NLR | **Decrease** | AFO-202 +N-163 |  |
| **Metabolic regulation** | HbA1c | **Decrease** | AFO-202 | <0.05 |
|  | Glycated Albumin |  | AFO-202 | <0.05 |
|  | T-Cho |  | AFO-202 +N-163 | <0.05 |
|  | LDL-Cho |  | AFO-202 +N-163 | <0.01 |
| **Coagulopathy prevention** | D-Dimer | **Decrease** | AFO-202 | <0.05 |
|  | Ferritin |  | AFO-202 +N-163 |  |
|  | Fibrinogen |  | AFO-202 +N-163 |  |
|  | Galectin-3 |  | AFO-202 +N-163 |  |
|  | CD11b | **Increase** | AFO-202 +N-163 |  |

## Discussion

With over a billion of people, or one quarter of the world’s population, affected with metabolic syndrome (MeTS) characterized by abdominal obesity, insulin resistance, hypertension, and hyperlipidaemia, MeTS has become a major health hazard in the modern world [20]. Consumption of high-calorie, low-fibre fast food and the decrease in physical activity combined with genetic predisposition fuel the rapid growth of this global epidemic. The cost of healthcare and loss of potential economic activity due to MeTS is in the trillions, and MeTS leads to other diseases such as chronic kidney disease, fatty liver disease, etc. [20]. Although lifestyle changes remain the primary modality of therapy, several drugs, including statins and anti-diabetic medications, are major agents used in therapy, which do little to treat the secondary symptoms and are associated with side effects [21, 22].

Furthermore, these therapeutic strategies separately address either glucotoxicity caused by abnormal and uncontrolled blood glucose levels or lipotoxicity due to a dysregulated lipid profile, but do not attempt to address the immune disruptions induced by MeTS [1, 4], which in turn enhances the development and progression of the associated chronic diseases. Advancing metabolic disruption, enhanced by aging-induced inflammatory disorders (inflammaging), leads to accumulation of lipids in the aging organs, coupled with immunosenescence [3] which further increases individuals’ risk of contracting infectious diseases. Importantly, the therapeutic strategies have a role after the onset of the disease, but they are not administered as a prophylaxis.

A continuous safe supplementation approach, which could serve as a prophylaxis before the onset of disease and an adjunct to existing treatments after disease onset, could be a holistic solution for which the *A. pullulans*’ novel strains-produced exopolysaccharide beta glucan-based biological response modifiers could be of potential use. The *A. pullulans* is a polyextremotolerant generalist black yeast belonging to the phylum Ascomycota, class Dothideomycete and order Dothideales having high levels of genetic recombination [23]. The AFO-202 strain of this black yeast produced beta glucan, having been documented to alleviate glucotoxicity and enhance immunity [8–10], when combined with the N-163 strain produced beta glucan has significant balancing effects on the lipid profile, anti-inflammatory and anti-fibrotic effects with immunomodulation [16–17], are further substantiated by the results of the present study.

In the present study in healthy Japanese men, AFO-202 has been shown to enhance the immune system, as observed from the increase in eosinophils and monocytes. A decrease in CRP observed in Group I (Figure 3C) with CRP being an acute phase reactant and known to increase rapidly with the onset of cell injury and inflammation shows that the AFO-202 beta glucan has anti-inflammatory and immune enhancement potential [24]. A significant increase in CD11b in the monocyte fraction was observed after 21 days of ingestion (Figure 6D). CD11b is expressed on monocytes, macrophages, dendritic cells, granulocytes, and NK cells and is an LPS receptor [25]. It is associated with the bacteriophagocytic activity of phagocytes. The increase in CD11b in the monocyte fraction of Group II after 21 days of consumption compared with before can be considered as a manifestation of immune activation by N-163 beta glucan. There was no change in IgG or IgM levels in the blood throughout the study period in both test groups, but an increase in IgA levels along with the decrease in D-dimer by AFO-202 and of galectin, fibrinogen, and ferritin by the combination of AFO-202 and N-163 beta glucan (Group II) offers evidence in favour of the combined approach for addressing immune associated coagulopathy-associated risks in diseases such as COVID-19, in which a hyperactivated immune response affects the clotting pathway [11]. The decrease in NLR with an increase in LCR and LeCR, all having been reported to be potential biomarkers of the underlying inflammation and hyperactivated immune response in COVID-19 [11], further substantiate the potent anti-inflammatory potential of these beta glucans. In terms of the secondary endpoints of glycaemic control and normalization of cholesterol levels, a longitudinal comparison showed a significant decrease in HbA1c and GA after 21 days of consumption in Group I and in T-Cho and LDL-Cho in Group II, suggesting that the synergistic intake of these beta glucans suppresses the increase in blood glucose level and lowers the cholesterol level, which could be useful in the context of metabolic disorders with underlying immune dysregulation, which again is a key factor associated with disease severity and mortality in COVID-19 [13], apart from application in management of MeTS and associated cardiac/cardiovascular abnormalities [26, 27].

Having illustrated the beneficial effects of the two strains of *A.pullulans* produced beta glucans, whose immune enhancement and immune modulation effects are complemented by their capability to balance metabolism, with potential prophylaxis and supplemental therapeutic approaches, two important arenas of further research are essential for arriving at a holistic understanding and enhancement of their benefits. One is the process of aging, against which all mechanisms must act, as aging is an inevitable phenomenon causing weakness of the whole human body, from the cellular to organ level, and age-related cumulative pathogenesis. The second essential component of research must be on the gut microbiota, also called the “second genome” [28], as their involvement and contribution to both metabolism balancing and immune modulation, besides neuronal implications for aging apoptosis, chronic micro-inflammation, and carcinogenesis [29], have been gaining strong evidence in the past decade. Immune weakness with aging having been known due to weakening hematopoietic cell and stem cell systems, a dent through which infections and aberrant / mutagenic cancer cells gain an entry to form a full-blown infection or cancer. With the earlier studies of immune cell enhancement in young healthy volunteers and elderly cancer patients [31] having been earlier proven as well with these beta glucans, a large population involvement to document the same would strengthen such findings. As beta glucans are known for their pre-biotic effects, and their beneficial effects could be proven to correlate with the gut microbiota, we should be able to see an amalgamation of all these and their mechanisms of interaction to explain technically their various potentials in terms of prevention, prophylaxis, and as a therapeutic adjunct for both communicable and non-communicable diseases.

It should be noted that this study was performed in healthy volunteers as an exploratory study, which warrants further validation in translational models designed for specific diseases to confirm the efficacy in specific pathogenesis and in human clinical studies in target illnesses. Although the study sample was small, this study paves the way for future research on the effects of these safe nutritional supplements in prophylaxis and prevention of disease in high-risk individuals, such as those with MeTS, as well as in infectious and non-infectious immune-metabolic dysfunction-associated diseases such as COVID-19.

## Conclusion

In summary, this study has demonstrated that the AFO-202 beta glucan is capable of eliciting beneficial effects in balancing blood glucose, alleviating glucotoxicity with immune activation, while a combination of AFO-202 and N-163 beta glucans has significant anti-inflammatory and lipid profile regulating potential, thereby alleviating lipotoxicity. Therefore, these beta glucans, being safe to consume, can be incorporated as a new beneficial adjunct therapy for individuals with developing and established MeTS. Future studies in subjects with relevant illnesses are warranted to evaluate these beta glucans’ application as agents for prophylaxis and management of fibrosis-induced non-communicable diseases such as fatty liver disease and immune hyperactivation-related morbidity and mortality, especially in communicable diseases such as COVID-19. Further elaborate research on evaluation of these beta glucans for their pre-biotic potentials in gut microbiota and related outcomes in managing chronic microinflammation, apoptotic mechanisms, and carcinogenesis could lead to evolution of knowledge and henceforth applications.

## Data Availability

All data generated or analyzed during this study are included in this manuscript

## Acknowledgements

The authors wish to acknowledge,

a. M/s CPCC Co. Ltd. and M/s Chiyoda Paramedical Care Clinic, Tokyo, Japan for their assistance in planning and execution of the entire study.
b. Mr. Yasunori Ikeue, Dr. Mitsuru Nagataki and Mr. Takashi Onaka, (Sophy Inc, Kochi, Japan), for necessary technical clarifications.
c. Dr. Kadalraja Raghavan, Research & Development Division, Sarvee Integra Pvt Ltd, Chennai, India and Dr. Vaddi Suryaprakash, Department of Urology, Yashoda Hospitals, Hyderabad, India for their technical inputs.
d. Mr. Yoshio Morozumi, Ms. Yoshiko Amikura of GN Corporation, Japan for their liaison assistance with the conduct of the study.
e. Loyola-ICAM College of Engineering and Technology (LICET) for their support to our research work.

